# A systematic review of dengue outbreak prediction models: current scenario and future directions

**DOI:** 10.1101/2022.07.06.22277291

**Authors:** Xing Yu Leung, Rakibul M. Islam, Mohammadmehdi Adhami, Dragan Ilic, Lara McDonald, Shanika Palawaththa, Basia Diug, Saif U. Munshi, M.D. Nazmul Karim

**Affiliations:** School of Public Health and Preventive Medicine, Monash University, Melbourne, Victoria, Australia; Department of Virology, Bangabandhu Sheikh Mujib Medical University, Dhaka, Bangladesh

## Abstract

Dengue is among the fastest-spreading vector-borne infectious disease, with outbreaks often overwhelm the health system and result in huge morbidity and mortality in its endemic populations in the absence of an efficient warning system. A large number of prediction models are currently in use globally. As such, this study aimed to systematically review the published literature that used quantitative models to predict dengue outbreaks and provide insights about the current practices. A systematic search was undertaken, using the Ovid MEDLINE, EMBASE, Scopus and Web of Science databases for published citations, without time or geographical restrictions. Study selection, data extraction and management process were devised in accordance with the ‘Checklist for Critical Appraisal and Data Extraction for Systematic Reviews of Prediction Modelling Studies’ (‘CHARMS’) framework. A total of 78 models were included in the review from 51 studies. Most models sourced climate (89.7%) and climate change (82.4%) data from agency reports and only 59.0% of the models adjusted for reporting time lag. All included models used climate predictors; 65.4% of them were built with only climate factors. Climate factors were used in combination with climate change factors (10.3%), both climate change and demographic factors (10.3%), vector factors (5.1%), and demographic factors (5.1%). Machine learning techniques were used for 38.5% of the models. Of these, random forest (20.0%), neural networks (23.3%) and ensemble models (13.3%) were notable. Among the statistical (61.5%) models, linear regression (20.8%), Poisson regression (18.8%), generalized additive models (16.7%) and time series/autoregressive models (18.8%) were notable. Around 24.4% of the models reported no validation at all and only 6.4% reported external validation. The reporting of methodology and model performance measures were inadequate in many of the existing prediction models. This review collates plausible predictors and methodological approaches, which will contribute to robust modelling in diverse settings and populations.

## Introduction

Dengue fever is one of the fastest-spreading mosquitos-borne disease primarily of tropical and subtropical regions and is caused by various dengue virus strains.^1,2^ Every year around 390 million people acquire the infection, of which around 96 million develop clinical manifestations.^3^ Although most infections are mild, dengue shock syndrome and dengue haemorrhagic fever are severe forms of infections and can be fatal.^4,5^ The case-fatality rate can be as high as 20% in the absence of prompt diagnosis and lack of specific antiviral drugs or vaccines,^6,7^ particularly in resource-limited settings. When the outbreak is particularly large, the influx of severe dengue cases can overwhelm the health system and prevent optimal care. Dengue also imposes an enormous societal and economic burden on many of the tropical countries where the disease is endemic.^8^ An accurate prediction of the size of the outbreak and trends in disease incidence early enough can limit further transmission,^5^ and is likely to facilitate planning the allocation of healthcare resources to meet demand during an outbreak.

Vector-borne pathogens characteristically demonstrate spatial heterogeneity - a result of spatial variation in vector habitat, climate patterns and subsequent human control actions.^9–11^ The interplay of human, climate and mosquito dynamics give rise to a complex system that determines the pattern of dengue transmission, which in turn influences the potential for outbreak.^12^ These relationships have been explored over the decades in the development of predictive models worldwide. Models vary widely in their purposes^13–15^ and settings.^16–21^ Many of these models excel at different tasks, however for a prediction model to be efficient, it requires a systematic, self-adaptive and generalizable framework capable of identifying weather and population susceptibility patterns across geographic regions. The scientific community has not yet agreed upon a model that provides the best prediction. The selection of predictors for the existing models is also quite heterogeneous. Some models rely solely on climate variables,^16^ some include vector characteristics^17,18^ others use population characteristics.^19-21^ A wide range of statistical techniques are used with varying degrees of accuracy and robustness among the existing models.^16–21^

Clarity in the documentation of the model development processes and model performance are essential for ensuring the robustness of the prediction,^22^ which is scarce as many of the existing models have not yet been systematically appraised. Given the disparate approaches, a focused synthesis and appraisal of the existing models, along with their building techniques and factor catchments, is required. Carefully establishing these details will provide the foundation for updating and developing robust models in future. This study aimed to systematically review all published literature that reported quantitative models to predict dengue outbreaks, revealing several shortcomings in the usage of real time primary predictive data and non-climatic predictors in the development of models, as well as inadequate reporting of techniques, model and performance measure validation.

## Methods

### Search strategy and selection criteria

This systematic review’s aim, search strategy and study selection process were devised in accordance with the seven items in the CHARMS framework.^23^ The review followed the Preferred Reporting Items for Systematic Review and Meta-Analysis (‘PRISMA’) guidelines,^24^ and was registered in PROSPERO (CRD42018102100).

A literature search was conducted from inception until July 2021 using the electronic databases of Ovid MEDLINE, Embase, Scopus and Web of Science to obtain the information on the statistical models for predicting the number of dengue cases based on climatic factors. Google Scholar and the bibliography of included papers were also searched. The search strategies were developed under the guidance of an information specialist from Monash University Library. For the purposes of this study, dengue fever or dengue haemorrhagic fever or dengue shock syndrome were considered as a single entity “dengue”. Search strategy included Medical Subject Headings (‘MeSH’) and keyword terms including “dengue”, “severe dengue,” “weather,” “climate change,” “model,” “predict,” and “forecast.” The detailed search strategy and history are presented in S1 Table.

The review included studies focused on (1) prognostic prediction models which aim to review models predicting future events, (2) models intended to inform public health divisions of future dengue outbreaks, (3) prediction model development studies without external validation, or with external validation in independent data, (4) people with dengue or dengue fever or dengue haemorrhagic fever, (5) the number of dengue cases based on climate and other factors, (6) models with no restrictions on the time span of prediction and (7) models to be used to predict the number of cases before outbreaks.

Peer-reviewed original articles that presented a model and were available as full-text articles were considered eligible if they focused on predicting the number of dengue cases or an outbreak based on number of dengue incidence. Articles that focused on updating previously developed models were only included if they presented an updated version of the model. Articles which dealt exclusively with dengue in international travellers, or which only analyse the correlation between climate parameters and dengue cases without presenting a prediction model were excluded. Furthermore, articles which used models for predicting the population of dengue vectors (e.g., *Aedes aegypti* or *Aedes albopictus*) as well as articles which only offer susceptible-infected-recovered modelling stochastic or transmission rates modelling were excluded. Articles which presented a model only dealing with spatial or temporal components of dengue risk were considered ineligible. Conference proceedings, book chapters, abstracts or letters were also excluded. Titles and abstracts of the retrieved articles were screened independently by two reviewers (RMI, MMA). Two review team members (LM, XYL) then retrieved the full text of those potentially eligible studies and independently assessed their eligibility. Disagreements were resolved by a third reviewer (MNK). A detailed study selection process is illustrated in the PRISMA flow diagram (Fig 1).^24^

**Figure 1.**
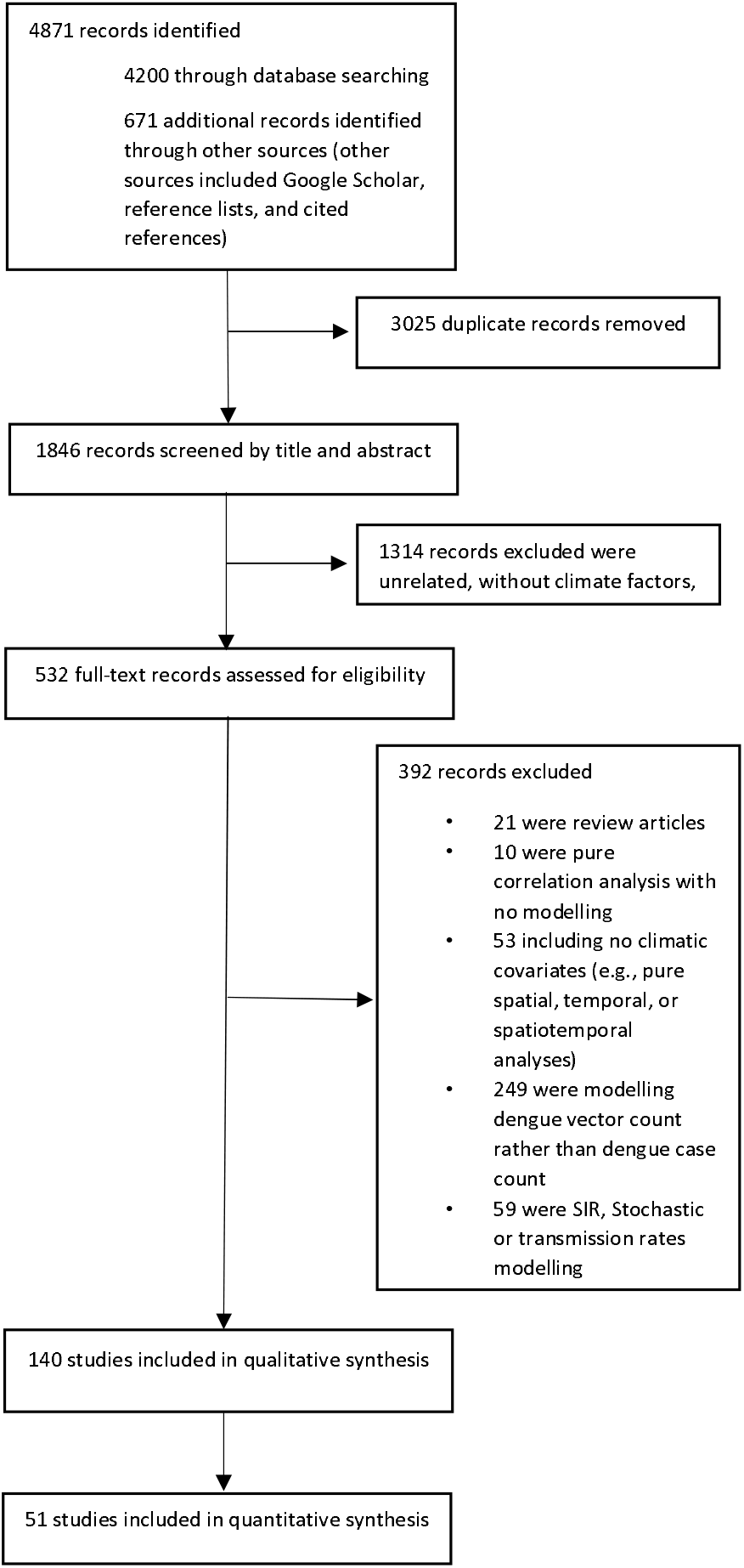
PRISMA flow diagram illustrating study selection process.

### Data analysis

Based on the data extraction fields of the CHARMS framework,^23^ a standardised table was developed to extract data from the selected studies for assessment of quality and evidence synthesis. The data extraction table consists of eleven domains, each with a specific item, that extract data from the reports of the primary forecasting model. Key information extracted from the included articles were period and geographical region, sources of data, outcomes to be predicted, modelling covariates variables, sample size, statistical techniques, model performances, model evaluation, and key findings. Each paper was independently reviewed by two reviewers (MMA, XYL) and discrepancies were resolved through discussion with each other or with a third reviewer (RMI) where necessary.

Extracted data from the selected studies were summarised and the key information about the methodological characteristics of these models were tabulated. Descriptive statistics were generated based on model characteristics and comparative methodological features such as outcome types, target population, data sources and predictor selection techniques. All statistical analyses were performed using Stata (version 16.0).

### Role of the funding source

There was no funding for this study.

## Results

The initial search yielded 4871 studies. After duplicates were removed, 1846 studies were screened for titles and abstracts. This led to 140 studies for full text review, and 51 that strictly met the inclusion criteria (Fig 1), 11 of these studies reported multiple models. A total of 78 models from 51 selected studies were identified (Table 1).^14,15,17–21,25–68^

**Table 1.**
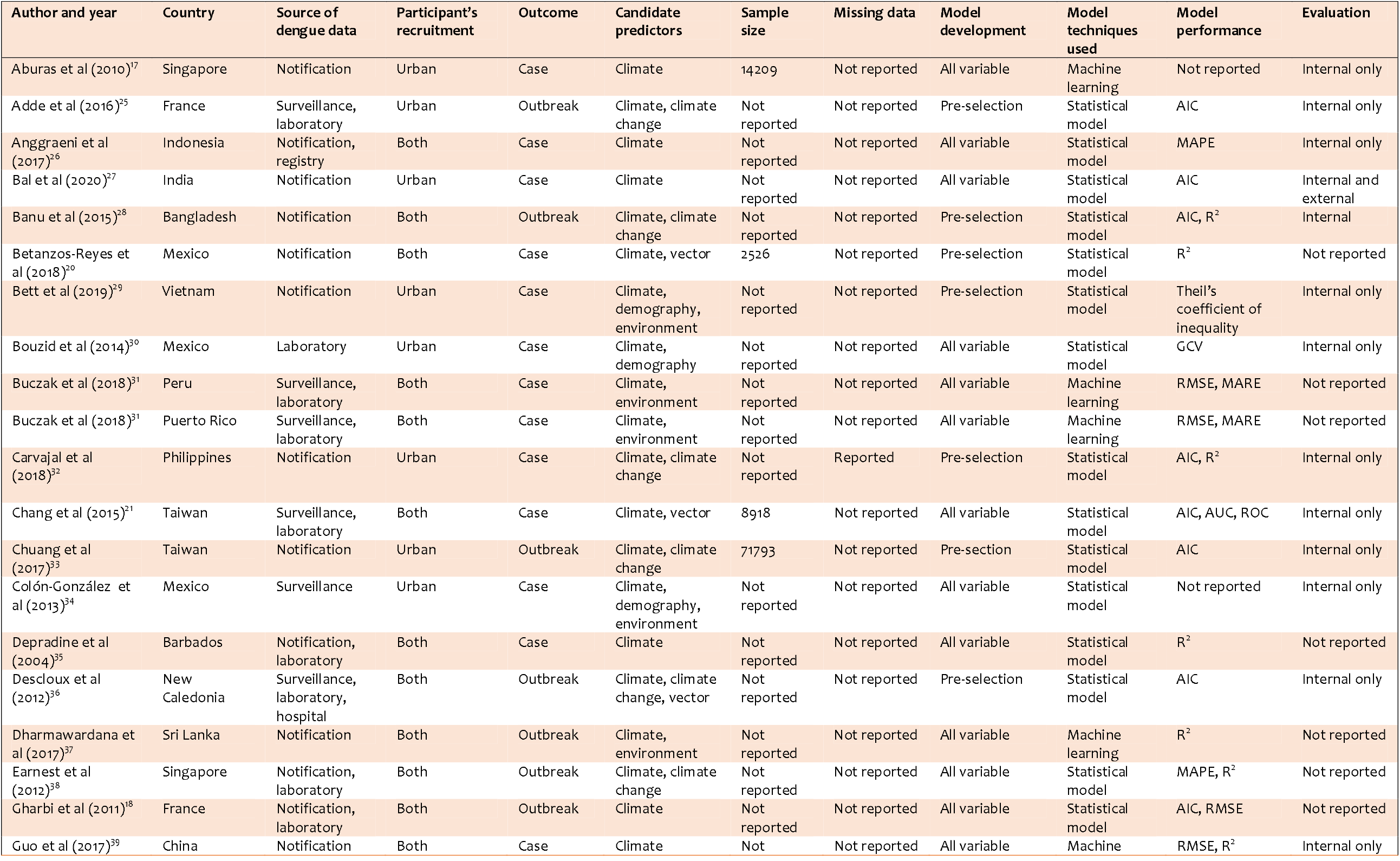
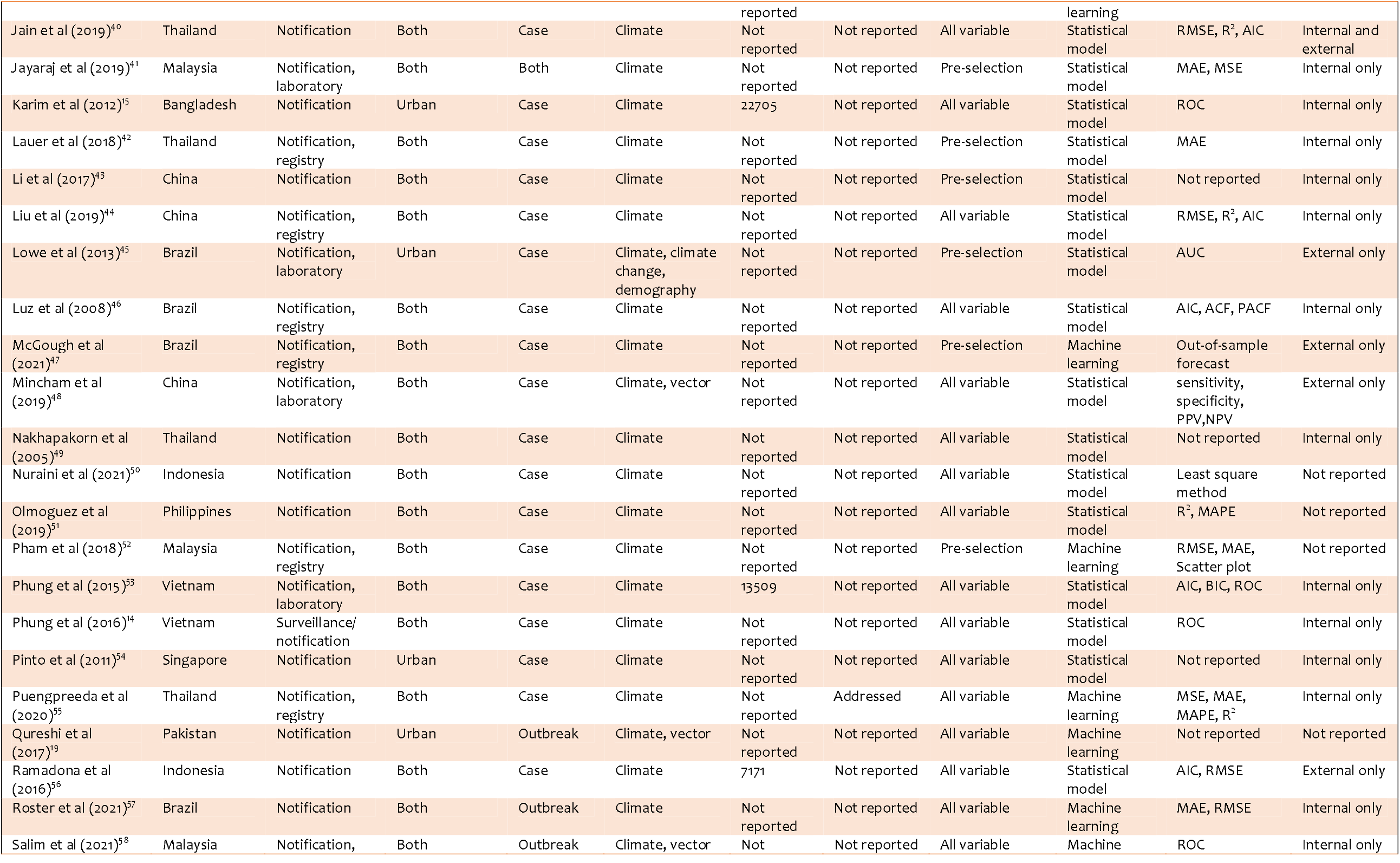
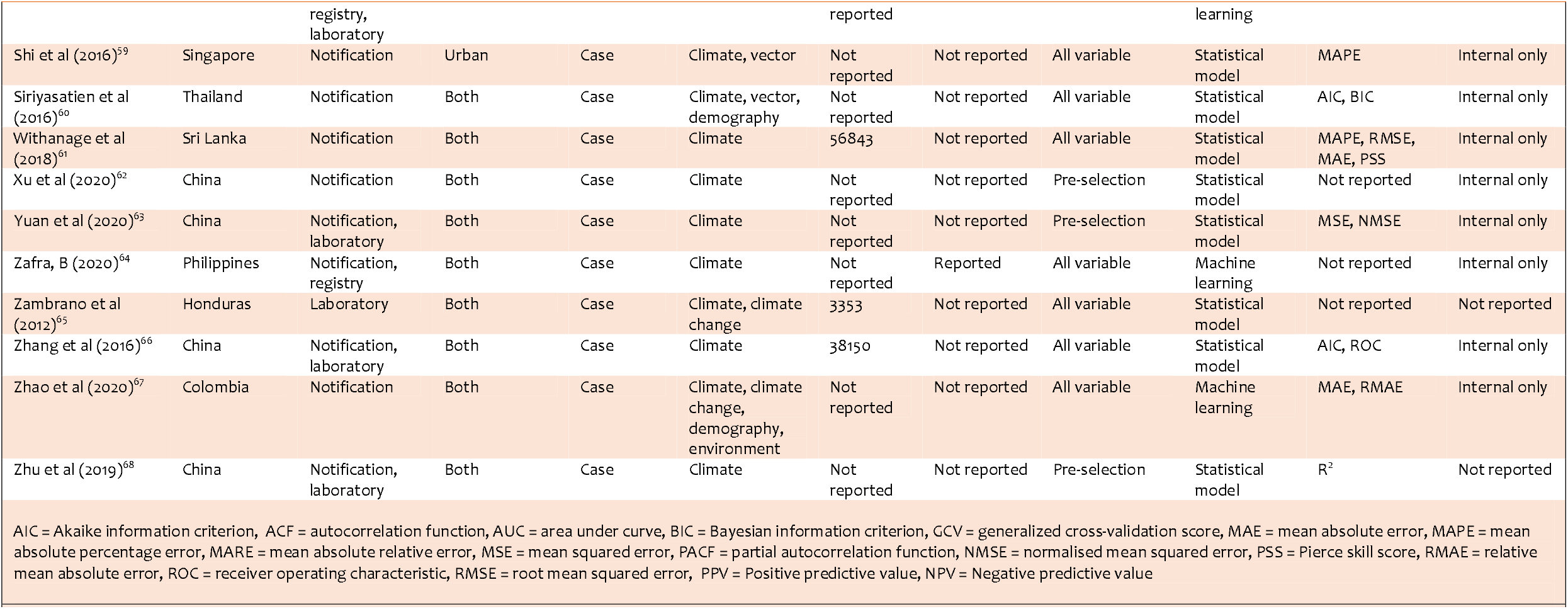
Characteristics of included predictive models.

Table 2 presents the sources of data used for modelling. Most of the models (93.6%) sourced dengue incidence data from surveillance and around a third used registry data, while another third also used hospital or laboratory data. While most (89.7%) of the models used climate data from government agency reports, only around 21.8% of the models used data from the meteorological stations in real-time. Climate change data was also sourced mostly (82.4%) from government agency reports, only 5.9% used international environmental agency data and 11.8% used local environmental agency report. Half (50.0%) of the models used vector data from entomological surveillance and a third used vector data from laboratory sources. Around 79.5% of the models were built based on the sample from general population, 20.5% used only urban samples. Around 42.9% of the models used monthly aggerate data, over a third (35.1%) used weekly aggregate data and 22.1% used daily aggregated data of the predictors. The majority (59.0%) of the models incorporated reporting time lag adjustment. Although 11.5% of the models addressed the missing data, 25.6% did not address the issue, while the majority (62.8%) did not specifically report the missing value. Around 76.9% of the models were intended to predict the number of dengue cases and 23.1% focused on predicting dengue outbreaks, based on predetermined case number threshold.

**Table 2.** Source of data used for modelling.

| Sources of model data | N (%) * |
| --- | --- |
| *Dengue data source (n=78)* |  |
| Surveillance/notification | 73 (93.6) |
| Disease registry | 25 (32.1) |
| Hospital/laboratory | 24 (30.8) |
| *Climate data source (n=78)* |  |
| Government agency report | 70 (89.7) |
| Meteorology station | 17 (21.8) |
| Research Institute/centre | 2 (2.6) |
| *Climate change data source (n=17)* |  |
| Government agency report | 14 (82.4) |
| International environmental agency | 1 (5.9) |
| Local environmental agency report | 2 (11.8) |
| *Vector data source (n=7)* |  |
| Surveillance/monitoring data | 3 (50.0) |
| Laboratory data | 2 (33.3) |
| Government agency report | 2 (33.3) |
| *Population source (n=78)* |  |
| General population | 62 (79.5) |
| Metropolitan | 16 (20.5) |
| *Data aggregation unit (n=77)* |  |
| Daily aggregate | 17 (22.1) |
| Weekly aggregate | 27 (35.1) |
| Monthly aggregate | 33 (42.9) |
| *Lag time adjusted in model (n=78)* |  |
| No | 32 (41.0) |
| Yes | 46 (59.0) |
| *Treatment of missing data (n=78)* |  |
| Yes | 9 (11.5) |
| No | 20 (25.6) |
| Not reported | 49 (62.8) |
| *Prediction outcome (n=78)* |  |
| Dengue case count | 60 (76.9) |
| Dengue outbreak | 18 (23.1) |
| * The percentages may not add up to 100 as studies obtained data from multiple sources |  |
| **Table 2. Source of data used for modelling** |  |

Table 3 presents the factors used for prediction models. All of the models included in the review used climate predictors in their model. Among the climate predictors: humidity (79.5%), temperature (98.7%) and rainfall (83.3%), were used in most models. Windspeed and direction (30.8%), precipitation (16.7%) and sunshine (11.5%) were among other notable climate factors. Overall, 21.8% of the models used climate change and environmental predictors. El Nino-Southern Oscillation (‘ENSO’), Southern Oscillation Index (‘SOI’), Oceanic Nino Index (‘ONI’), hydric balance and vegetation index were among the key climate change predictors. Vector-related predictors were included in 9.0% of models, and the key vector related predictors were container index, Breteau index, adult productivity index, breeding percentage and mosquito infection rate. Demographic predictors were included in 17.9% of models, and key demographic predictors were, population size, population density, area under the urban settlement, access to piped water, education coverage and GDP per capita (Table 3).

**Table 3.** Factors appeared as predictors in the prediction models.

| Climate factors (100.0%) | Climate change and environmental factors (21.8%) | Entomological (Vector) factors (9.0%) | Demographic factors (17.9%) |
| --- | --- | --- | --- |
| Temperature (98.7%) | El Nino-Southern Oscillation (ENSO) | Ades albopictus count | Population size |
| Minimum temperature | Southern Oscillation Index (SOI) | Container index | Population density |
| Mean temperature | Oceanic Nino Index (ONI) | Ades aegypti index | Access to piped water |
| Maximum temperature | Gini Index | Breteau index | Education coverage |
| Rainfall (83.3%) | Potential evapotranspiration | Adult productivity index | GDP per capita |
| Average rainfall | Azores high sea-level pressure | Weekly egg count in ovitrap | Area under urban settlement |
| Accumulated rainfall | Dipole mode index | Breeding percentage |  |
| Number of rainy days | Hydric balance | Mosquito infection rate |  |
| Humidity (79.5%) | Vegetation Index | Minimum infection rate |  |
| Relative humidity | Equatorial Pacific Ocean surface temperature | Per man hour density (PMHD) |  |
| Absolute humidity |  |  |  |
| Sunshine (11.5%) |  |  |  |
| Sunshine duration |  |  |  |
| Insolation |  |  |  |
| Windspeed & direction (30.8%) |  |  |  |
| Precipitation (16.7%) |  |  |  |
| Evaporation (9.0%) |  |  |  |
| Atmospheric pressure (2.6%) |  |  |  |
| * Figures in the parenthesis denotes the percentage of models included the predictor in the model and percentages may not add up to 100 as models used multiple categories of predictors in combination |  |  |  |
| **Table 3. Factors appeared as predictors in the prediction models** |  |  |  |

The combination of the predictors used in the model are depicted in Figure 2. While majority (65.4%) of the models were built solely on climate predictors, none of the models used the combination of all four (climate, climate change, vector and demography) categories of predictors. The combination of climate, climate change and demographic predictors was used in 10.3% of the models and the combination of climate and climate change predictors were used in 10.3% models. Among other notable combinations were, climate and vector predictors (5.1%) and climate and demographic predictor (5.1%).

**Figure 2.**
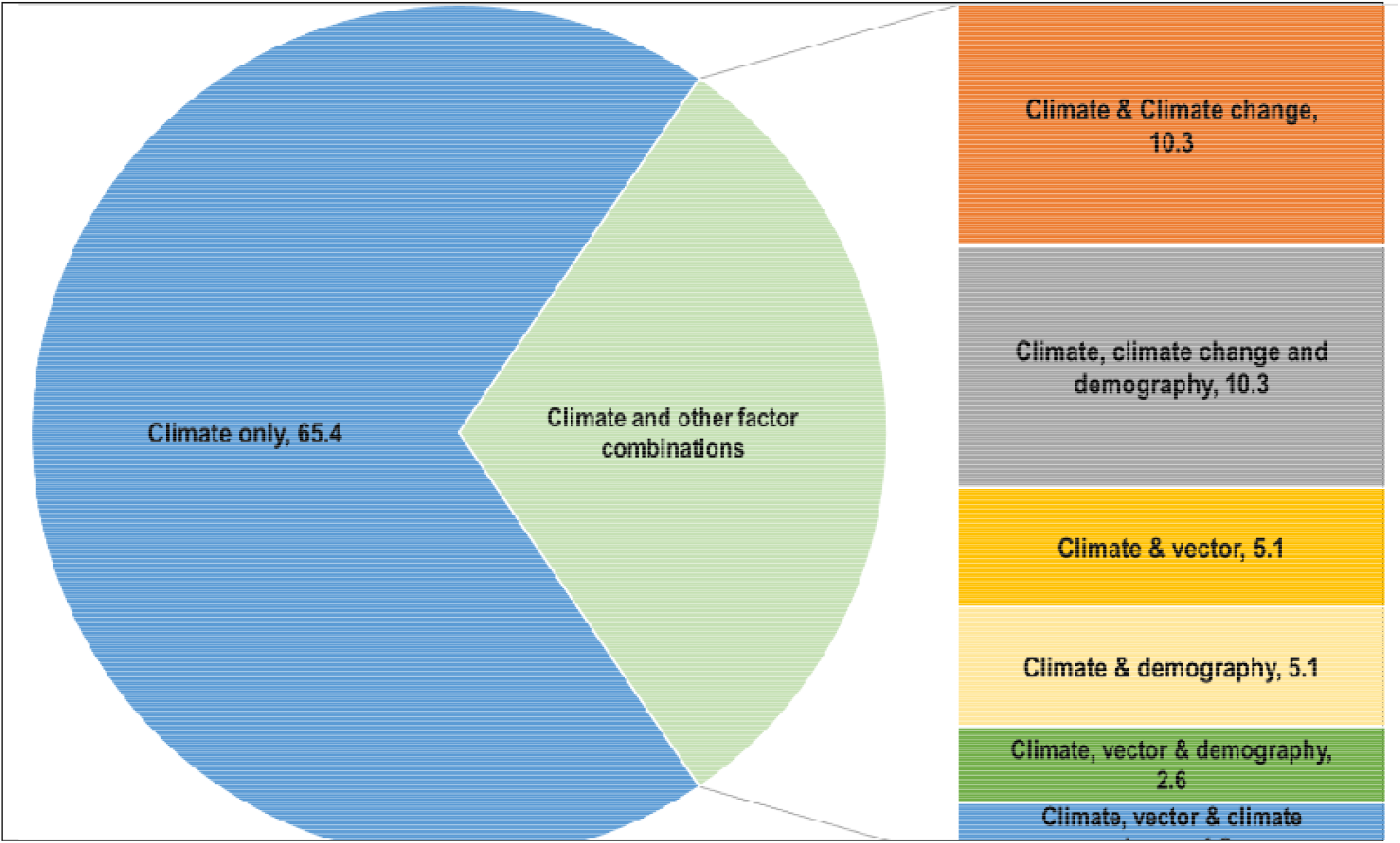
Combinations of predictors used in the prediction models.

Table 4 summarises the statistical methods adopted by the prediction models. Modelling techniques were broadly categorised under two genres, statistical models (61.5%) and machine learning (38.5%). The statistical models were broadly comprised of linear regression models (20.8%), time series/autoregressive models (18.8%), Poisson regression models (18.8%) and generalized additive models (16.7%). Neural networks models (23.3%), random forest models (20.0%), and ensemble models (13.3%) were types of machine learning models used.

**Table 4.**
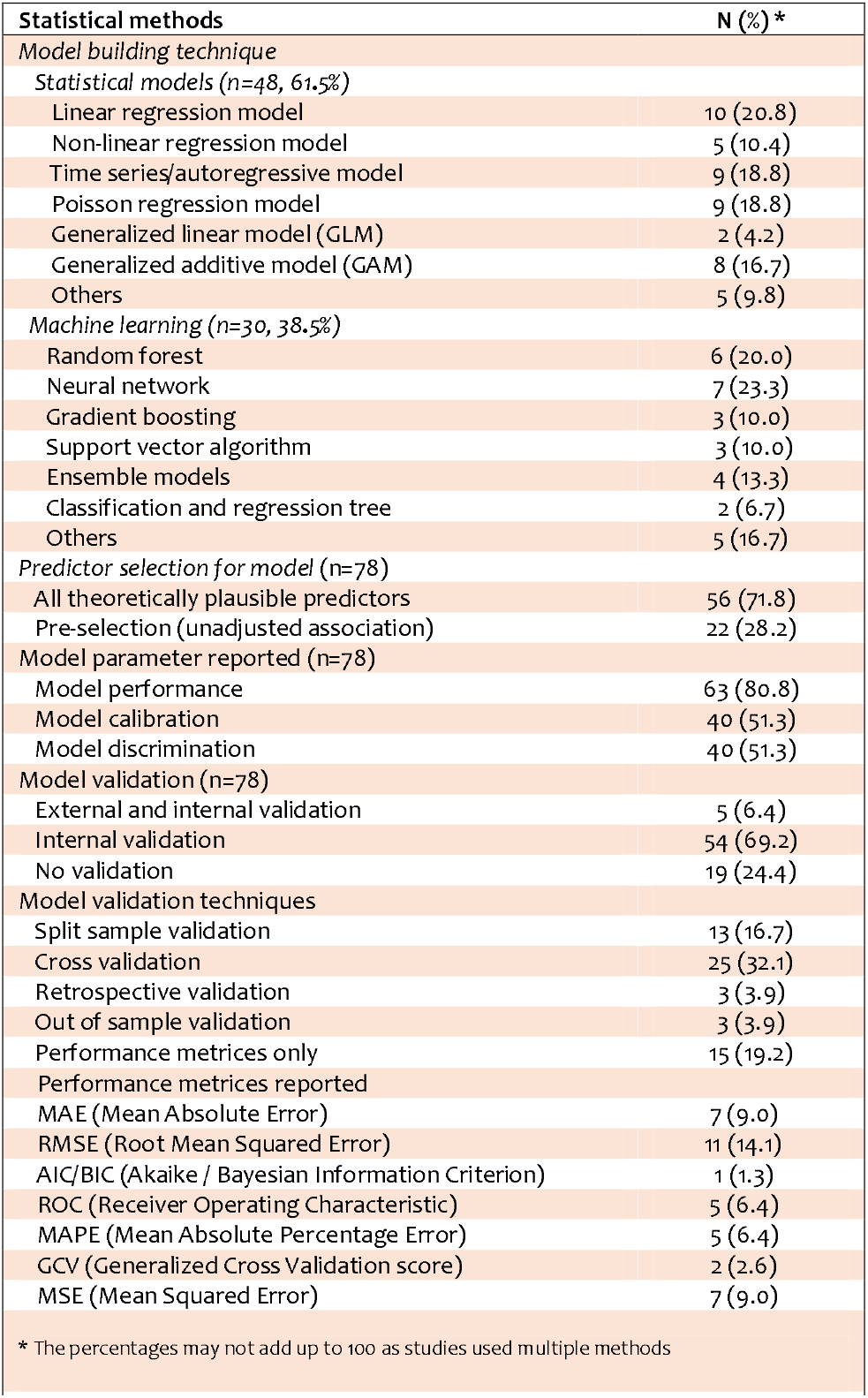
Statistical methods used among models (n=78)

All theoretically plausible predictors were considered as candidate predictors in 71.8% of models and pre-selection of predictors based on unadjusted association with the model outcome was considered in 28.2% of models. Reporting of essential modelling techniques was heterogeneous – 80.8% of models reported model performance, 51.3% reported model calibration and 51.3% reported model discrimination.

Of these models, most (69.2%) reported the internal validation alone, only 6.4% reported both internal and external validation and 24.4% reported no validation at all. The validation techniques included: split sample validation (development and validation) (16.7%), cross validation, which involves resampling of the derivation sample (32.1%) and performance metrics (19.2%). Among the performance metrices, Root Mean Squared Error (‘RMSE’) (14.1%), Mean Squared Error (‘MSE’) (9.0%), Mean Absolute Percentage Error (‘MAPE’) (6.4%), and Receiver Operating Characteristic (‘ROC’) (6.4%) were notable.

## Discussion

This systematic review evaluated 78 dengue outbreak prediction models from 51 studies, predicting the number of dengue cases or outbreaks from a variety of settings and populations. Our review identified, three major area of inadequacy in the current modelling practices. Firstly, use of secondary predictor data—acquired from reports—were quite prevalent among models. Secondly, the majority of the models ignored non-climatic variables while the model was being developed, and thus failed to capture a holistic view of dengue development in the prediction process. Lastly, inadequacy in the reporting of methodology, model validation and performance measures were quite prevalent in the existing prediction models. One positive aspect seen in the current modelling practice is the shift toward robust modelling technique, such as use of machine learning algorithm and autoregressive time series techniques.

While effective treatments and prevention measures are still being developed, an early warning system for an epidemic has the potential to reduce the toll of severe disease on the health system and population.^69^ Considering dengue is a mosquito-borne disease, the majority of outbreak prediction models focus on climate dependency of mosquito breeding and dengue transmission.^4–7^ While many models have been successful in predicting relative cases of dengue in real settings, incorrect prediction results have been observed in several included studies. For example, a model by Adde et al.^25^ was unable to forecast a dengue outbreak in 2001–2005 with the use of climate data from 1991–2000. One of the potential reasons for their inaccurate prediction is the geo-spatial variation of climate and environment within regions. In their study, the decision to include vastly heterogenous geographical areas led to variation in model prediction, which— be due to exclusion of non-climatic factors—may be the explanation of the poor performance in many of the earlier prediction models could. An increasing number of later models appears to incorporate a wide range of vector parameters as well as demographic parameters. Chang et al pointed out that, entomological (vector) factor combined with other meteorological (climate) factors, have better prediction performance, and their prediction accuracy is often higher than that of climate predictors alone. ^21^

For dengue incidence data, the majority of the models relied on reports from government organizations based on notifiable data. Notification involves passive surveillance, where there is potential for systematic underreporting along with varying time lag. Modelling with data from active surveillance or real-time study may minimize such limitations. A considerable number of models did not consider the time lag affecting the prediction, which may be responsible for possible delays in weather affecting mosquito vectors and subsequently viruses. Due to the nature of dengue disease dynamics, failure to address time lag in model development is likely to affect prediction accuracy. Critical points in the natural history of disease timeline those may generate time lag may start with mosquito development, and subsequently also during acquisition and amplification virus in mosquitoes, mosquito host behaviour (i.e. biting and feeding pattern) and the incubation period of the virus in the human body.^12,41^ Some studies have found a positive correlation between climate variables with time-lags at several points in the natural history of disease timeline.^41,45^ Therefore, the adjustment for the time lags while predicting dengue is indispensable, especially when meteorological data is used.^12^

The majority of included models were built on conventional regression techniques. According to recent literature, the time series technique is particularly considered effective in predicting the highly auto-correlated nature of dengue infection.^70,71^ Machine learning techniques are gaining increasing popularity, with around 30% 0f the included studies using such techniques, particularly prevalent among the recently developed models. Batista et al. confirmed superiority with ML techniques demonstrating a lower error rate compared to the conventional statistics-based model in predicting dengue cases. In the age of big data, this technique can leverage data availability and in addition to being non-parametric in nature, can also provide some leeway in terms of strict assumption.^72^ Random forest, neural networks, gradient boosting and support vector algorithms are notable subsets of machine learning algorithms, which have made significant contributions to several areas of public health, particularly in the forecasting of infectious diseases like malaria^73^ and COVID-19^74^, and may have similar utility for making dengue outbreak predictions.

In the modelling process, generating an algorithm or equation is only part of the process. It is not complete unless its performance has been assessed considering discrimination^75^ and calibration^76^, both internally and using the population outside of what it is developed from, respectively. Among the existing models examined, reporting of the discrimination and calibration is very low. Without knowledge of model performance through validation in both source population and population other than where it was developed, objective evaluation of models is difficult.^77^ In a substantial proportion of the models, those reported validation, the original dataset was randomly split into the development and validation subset.

Although this approach is widely used in many model validation settings, there are some setbacks when used with smaller operational sample sizes. Such splitting of the sample not only creates two smaller data sets that are similar but also increases the possibility of overfitting, with apparent performance. Therefore, for small data sets, using a split sample approach increases the risk of bias, while for large datasets there is no additional benefit.^78^ In terms of internal validation, bootstrapping is generally considered to be the preferred internal validation method in predictive models because it provides a contraction factor to adjust the estimated regression coefficient and the apparent model performance of such overfitting.^79^ Interestingly, bootstrapping was not used in any of the models in included studies, instead cross-validation technique was adopted in most of them. External validation, on the other hand, was used only in very few included studies. This is despite the fact that external validation is considered pivotal to model development and a key indicator of model performance through highlighting applicability to participants, centres, regions or environments.^23^ The external validation is particularly essential for model redevelopment, where the original model is adjusted, updated, or recalibrated based on validation data to improve performance.^80^ This update may include adjusting the baseline risk (interception or hazard) of the original model, adjusting the weight or regression coefficient of the predictor, adding new predictors, or removing existing predictors from the model.

This review has a number of strengths – specifically, the use of the CHARMS checklist,^23^ designed for the assessment of the applicability of the prediction models. In addition, inclusion and exclusion criteria were strictly followed, and database searches were conducted by an expert librarian. However, there are a few limitations of the review – the models in this review are not explicitly rated based on quality or performance due to the lack of accepted criteria for rating the quality of forecasting models. In addition, although calibration was reported in several studies, calibration measures lack clarification, which may impact the overall evaluation of the model performance. The model performance could not be compared across methodological approaches in quantitative synthesis because of a lack of model performance data, and those that did provide data are mostly generated from internal validation data which may result in overfitting.

## Conclusion

In conclusion, failure to use of real time primary predictor data, failing to incorporate non-climatic parameters as predictor and insufficient reporting of model development techniques, model validation and performance measure were the major inadequacies identified in the current modelling practice. The paradigm shift towards robust modelling techniques, such as the use of machine learning algorithms and autoregressive time series, is a significant positive trend in contemporary model practices. The findings of this review have the potential to lay the groundwork for improved modelling practices in the future. These findings will contribute to robust modelling in different settings and populations and have important implications for the planning and decision-making process for early dengue intervention and prevention.

## Data Availability

All relevant data are within the manuscript and its Supporting Information files.

## Acknowledgements

MNK, SM, and DI were responsible for the study concept and design. RMI, MMA, LM, SP, and XYL extracted and validated the data. MNK conducted the statistical analyses. RMI, BD, SP and XYL interpreted the data. MNK, RMI, SP and XYL drafted and revised the versions the manuscript. All authors had full access to the data in the study. All authors revised and critically analysed the manuscript for important intellectual content.

## Conflict of interest

The authors declare that they have no conflicts of interest.

## Data Availability Statement

Available upon request.

## Supporting Information Captions

**S1 Table.** Search strategy for OVID Medline, as performed in July 2021.

PRISMA 2020 Checklist
| Section and Topic | Item # | Checklist item | Location where item is reported |
| --- | --- | --- | --- |
| **TITLE** |  |  |  |
| Title | 1 | Identify the report as a systematic review. | Pg 1 – Title “A systematic review of dengue outbreak prediction models: current scenario and future directions” |
| **ABSTRACT** |  |  |  |
| Abstract | 2 | See the PRISMA 2020 for Abstracts checklist. | Pg 2 |
| **INTRODUCTION** |  |  |  |
| Rationale | 3 | Describe the rationale for the review in the context of existing knowledge. | Pg 3, lines 50 – 52<br>“Prior to this study, no systematic reviews or meta-analyses have examined the existing models based on their development methodology, predictor variables used and model performance.” |
| Objectives | 4 | Provide an explicit statement of the objective(s) or question(s) the review addresses. | Pg 5, lines 89-91<br>“This study aimed is to systematically review all published literature that reported quantitative models to predict dengue outbreaks” |
| **METHODS** |  |  |  |
| Eligibility criteria | 5 | Specify the inclusion and exclusion criteria for the review and how studies were grouped for the syntheses. | Pg 6, lines 110 – 115<br>“The review included studies focused on (1) prognostic prediction models which aim to review models predicting future events, (2) models intended to inform public health divisions of future dengue outbreaks, (3) prediction model development studies without external validation, or with external validation in independent data, (4) people with dengue or dengue fever or dengue haemorrhagic fever, (5) the number of dengue cases based on climate and other factors, (6) models with no restrictions on the time span of prediction and (7) models to be used to predict the number of cases before outbreaks.” |
| Information sources | 6 | Specify all databases, registers, websites, organisations, reference lists and other sources searched or consulted to identify studies. Specify the date when each source was last searched or consulted. | Pg 6, lines 101 – 104<br>“A literature search was conducted from inception until July 2021 using the electronic databases of Ovid MEDLINE, Embase, Scopus and Web of Science to obtain the information on the statistical models for predicting the number of dengue cases based on climatic factors. Google Scholar and the bibliography of included papers were also searched” |

PRISMA 2020 Checklist
| Section and Topic | Item # | Checklist item | Location where item is reported |
| --- | --- | --- | --- |
| Search strategy | 7 | Present the full search strategies for all databases, registers and websites, including any filters and limits used. | Pg 32, S1 Table. Search strategy |
| Selection process | 8 | Specify the methods used to decide whether a study met the inclusion criteria of the review, including how many reviewers screened each record and each report retrieved, whether they worked independently, and if applicable, details of automation tools used in the process. | Pg 7, lines 125 – 128<br>“Titles and abstracts of the retrieved articles were screened independently by two reviewers (RMI, MMA). Two review team members (LM, XYL) then retrieved the full text of those potentially eligible studies and independently assessed their eligibility. Disagreements were resolved by a third reviewer (MNK).” |
| Data collection process | 9 | Specify the methods used to collect data from reports, including how many reviewers collected data from each report, whether they worked independently, any processes for obtaining or confirming data from study investigators, and if applicable, details of automation tools used in the process. | Pg 9, lines 142 – 144<br>“Each paper was independently reviewed by two reviewers (MMA, XYL) and discrepancies were resolved through discussion with each other or with a third reviewer (RMI) where necessary.” |
| Data items | 10a | List and define all outcomes for which data were sought. Specify whether all results that were compatible with each outcome domain in each study were sought (e.g. for all measures, time points, analyses), and if not, the methods used to decide which results to collect. | Pg 9, lines 140 – 142<br>“Key information extracted from the included articles were period and geographical region, sources of data, outcomes to be predicted, modelling covariates variables, sample size, statistical techniques, model performances, model evaluation, and key findings” |
|  | 10b | List and define all other variables for which data were sought (e.g. participant and intervention characteristics, funding sources). Describe any assumptions made about any missing or unclear information. | Pg 9, lines 142 – 143<br>“Information regarding handling and/or reporting of missing data was also extracted.” |
| Study risk of bias assessment | 11 | Specify the methods used to assess risk of bias in the included studies, including details of the tool(s) used, how many reviewers assessed each study and whether they worked independently, and if applicable, details of automation tools used in the process. | This study did not pool data for meta-analysis, but instead was a largely qualitative appraisal focusing on model characteristics. Therefore, risk of bias assessment was not performed – the CHARMS framework was considered instead which by itself is a measure of quality. |
| Effect measures | 12 | Specify for each outcome the effect measure(s) (e.g. risk ratio, mean difference) used in the synthesis or presentation of results. | Model characteristics were primary focus of our largely qualitative appraisal |
|  | 13a | Describe the processes used to decide which studies were eligible for each synthesis (e.g. tabulating the study intervention characteristics and comparing against the planned groups for each synthesis (item #5)). | Pg 7, lines 125 – 129<br>“Titles and abstracts of the retrieved articles were screened independently by two reviewers (RMI, MMA). Two review team members (LM, XYL) then retrieved the full text of those potentially |

PRISMA 2020 Checklist
| Section and Topic | Item # | Checklist item | Location where item is reported |
| --- | --- | --- | --- |
|  |  |  | eligible studies and independently assessed their eligibility. Disagreements were resolved by a third reviewer (MNK). A detailed study selection process is illustrated in the PRISMA flow diagram (Fig 1)." |
|  | 13b | Describe any methods required to prepare the data for presentation or synthesis, such as handling of missing summary statistics, or data conversions. | Raw data extracted and included as per Table 1 |
|  | 13c | Describe any methods used to tabulate or visually display results of individual studies and syntheses. | N/A |
|  | 13d | Describe any methods used to synthesize results and provide a rationale for the choice(s). If meta-analysis was performed, describe the model(s), method(s) to identify the presence and extent of statistical heterogeneity, and software package(s) used. | Pg 9, lines 137 – 149<br>"Based on the data extraction fields of the CHARMS framework,<sup>23</sup> a standardised table was developed to extract data from the selected studies for assessment of quality and evidence synthesis. The data extraction table consists of eleven domains, each with a specific item, that extract data from the reports of the primary forecasting model. Key information extracted from the included articles were period and geographical region, sources of data, outcomes to be predicted, modelling covariates variables, sample size, statistical techniques, model performances, model evaluation, and key findings. Each paper was independently reviewed by two reviewers (MMA, XYL) and discrepancies were resolved through discussion with each other or with a third reviewer (RMI) where necessary.<br>Extracted data from the selected studies were summarised and the key information about the methodological characteristics of these models were tabulated. Descriptive statistics were generated based on model characteristics and comparative methodological features such as outcome types, target population, data sources and predictor selection techniques. All statistical analyses were performed using Stata (version 16.0)." |
|  | 13e | Describe any methods used to explore possible causes of heterogeneity among study results (e.g. subgroup analysis, meta-regression). | This study did not pool data for meta-analysis, but instead was a largely qualitative appraisal focusing on model characteristics. Therefore, heterogeneity was not considered to be of great relevance to the study aims. |
|  | 13f | Describe any sensitivity analyses conducted to assess robustness of the synthesized results. | This study did not pool data for meta-analysis, but instead was a largely qualitative appraisal |

PRISMA 2020 Checklist
| Section and Topic | Item # | Checklist item | Location where item is reported |
| --- | --- | --- | --- |
|  |  |  | focusing on model characteristics. Therefore, sensitivity analyses were not considered to be of great relevance to achieving our study aims. |
| Reporting bias assessment | 14 | Describe any methods used to assess risk of bias due to missing results in a synthesis (arising from reporting biases). | This study did not pool data for meta-analysis, but instead was a largely qualitative appraisal focusing on model characteristics. Therefore, risk of bias assessment was not performed – the CHARMS framework was utilised instead. |
| Certainty assessment | 15 | Describe any methods used to assess certainty (or confidence) in the body of evidence for an outcome. | Analysis was largely narrative with some summative statistics; meta-analysis and certainty assessments were not performed. |
| **RESULTS** |  |  |  |
| Study selection | 16a | Describe the results of the search and selection process, from the number of records identified in the search to the number of studies included in the review, ideally using a flow diagram. | Pg 8.<br>“Fig 1. PRISMA flow diagram illustrating study selection process” |
|  | 16b | Cite studies that might appear to meet the inclusion criteria, but which were excluded, and explain why they were excluded. | Pg. 8<br>As explained in Fig 1 |
| Study characteristics | 17 | Cite each included study and present its characteristics. | Pg 11 – 13<br>“Table 1: Studies of predictive modellings meeting inclusion criteria” |
| Risk of bias in studies | 18 | Present assessments of risk of bias for each included study. | This study did not pool data for meta-analysis, but instead was a largely qualitative appraisal focusing on model characteristics. Therefore, risk of bias assessment was not performed – the CHARMS framework was considered instead which by itself is a measure of quality. |
| Results of individual studies | 19 | For all outcomes, present, for each study: (a) summary statistics for each group (where appropriate) and (b) an effect estimate and its precision (e.g. confidence/credible interval), ideally using structured tables or plots. | Pg 15/16/18<br>“Table 2. Source of data used for modelling”,<br>“Table 3. Factors appeared as predictors in the prediction models”<br>“Table 4. Statistical methods used among models (n=78)” |
| Results of syntheses | 20a | For each synthesis, briefly summarise the characteristics and risk of bias among contributing studies. | This study did not pool data for meta-analysis, but instead was a largely qualitative appraisal focusing on model characteristics. Therefore, risk of bias assessment was not performed – the |

PRISMA 2020 Checklist
| Section and Topic | Item # | Checklist item | Location where item is reported |
| --- | --- | --- | --- |
|  |  |  | CHARMS framework was utilised instead. |
|  | 20b | Present results of all statistical syntheses conducted. If meta-analysis was done, present for each the summary estimate and its precision (e.g. confidence/credible interval) and measures of statistical heterogeneity. If comparing groups, describe the direction of the effect. | Pg 11 – 13<br>Tabulated in Table 1 |
|  | 20c | Present results of all investigations of possible causes of heterogeneity among study results. | This study did not pool data for meta-analysis, but instead was a largely qualitative appraisal focusing on model characteristics. Therefore, heterogeneity was not considered to be of great relevance to the study aims. |
|  | 20d | Present results of all sensitivity analyses conducted to assess the robustness of the synthesized results. | This study did not pool data for meta-analysis, but instead was a largely qualitative appraisal focusing on model characteristics. Therefore, sensitivity analyses were not considered to be of great relevance to achieving our study aims. |
| Reporting biases | 21 | Present assessments of risk of bias due to missing results (arising from reporting biases) for each synthesis assessed. | This study did not pool data for meta-analysis, but instead was a largely qualitative appraisal focusing on model characteristics. Therefore, risk of bias assessment was not performed – the CHARMS framework was considered instead which by itself is a measure of quality. |
| Certainty of evidence | 22 | Present assessments of certainty (or confidence) in the body of evidence for each outcome assessed. | Analysis was largely narrative with some summative statistics; meta-analysis and certainty assessments were not performed. |
| **DISCUSSION** |  |  |  |
| Discussion | 23a | Provide a general interpretation of the results in the context of other evidence. | Pg 20, lines 230 – 237<br>“Our review identified, three major area of inadequacy in the current modelling practices. Firstly, use of secondary predictor data—acquired from reports—were quite prevalent among models. Secondly, the majority of the models ignored non-climatic variables while the model was being developed, and thus failed to capture a holistic view of dengue development in the prediction process. Lastly, inadequacy in the reporting of methodology, model validation and performance measures were quite prevalent in the existing prediction models. One positive aspect seen in the current modelling practice is the shift toward robust modelling technique, such as use of machine learning algorithm and autoregressive time series techniques.” |
|  | 23b | Discuss any limitations of the evidence included in the review. | Pg 22, lines 300 – 306 |

PRISMA 2020 Checklist
| Section and Topic | Item # | Checklist item | Location where item is reported |
| --- | --- | --- | --- |
|  |  |  | “However, there are a few limitations of the review – the models in this review are not explicitly rated based on quality or performance due to the lack of accepted criteria for rating the quality of forecasting models. In addition, although calibration was reported in several studies, calibration measures lack clarification, which may impact the overall evaluation of the model performance. The model performance could not be compared across methodological approaches in quantitative synthesis because of a lack of model performance data, and those that did provide data are mostly generated from internal validation data which may result in overfitting.” |
|  | 23c | Discuss any limitations of the review processes used. | Pg 22, lines 301<br>“...the models in this review are not explicitly rated based on quality or performance” |
|  | 23d | Discuss implications of the results for practice, policy, and future research. | Pg 23, lines 314 – 317<br>“The findings of this review have the potential to lay the groundwork for improved modelling practices in the future. These findings will contribute to robust modelling in different settings and populations and have important implications for the planning and decision-making process for early dengue intervention and prevention.” |
| **OTHER INFORMATION** |  |  |  |
| Registration and protocol | 24a | Provide registration information for the review, including register name and registration number, or state that the review was not registered. | Pg 6, lines 99 – 100<br>“PROSPERO (CRD42018102100).” |
|  | 24b | Indicate where the review protocol can be accessed, or state that a protocol was not prepared. | Protocol accessible via PROSPERO (CRD42018102100) |
|  | 24c | Describe and explain any amendments to information provided at registration or in the protocol. | N/A |
| Support | 25 | Describe sources of financial or non-financial support for the review, and the role of the funders or sponsors in the review. | Pg 9, line 152<br>“There was no funding for this study.” |
| Competing interests | 26 | Declare any competing interests of review authors. | Pg 23, lines 323 – 324<br>“Conflict of interest<br>The authors declare that they have no conflicts of interest.” |

PRISMA 2020 Checklist
| Section and Topic | Item # | Checklist item | Location where item is reported |
| --- | --- | --- | --- |
| Availability of data, code and other materials | 27 | Report which of the following are publicly available and where they can be found: template data collection forms; data extracted from included studies; data used for all analyses; analytic code; any other materials used in the review. | Pg 23, lines 325 – 326<br>“Data Availability Statement<br>Available upon request.” |

*From:* Page MJ, McKenzie JE, Bossuyt PM, Boutron I, Hoffmann TC, Mulrow CD, et al. The PRISMA 2020 statement: an updated guideline for reporting systematic reviews. BMJ 2021;372:n71. doi: 10.1136/bmj.n71
For more information, visit: http://www.prisma-statement.org/

## Notes

### Competing Interest Statement

The authors have declared no competing interest.

### Funding Statement

The author(s) received no specific funding for this work.

