## Supplementary material for "A systematic review of dengue outbreak prediction models: current scenario and future directions": S1 Table

| **#** | **Searches** | **Results** |
| --- | --- | --- |
| 1 | Temperature/ | 223602 |
| 2 | temperature.mp. | 591394 |
| 3 | Weather/ | 9093 |
| 4 | weather.mp. | 17526 |
| 5 | Humidity/ | 15068 |
| 6 | humidity.mp. | 27232 |
| 7 | Rain/ | 9252 |
| 8 | rain*.mp. | 37542 |
| 9 | Wind/ | 3839 |
| 10 | wind.mp. | 11248 |
| 11 | Air Movements/ | 3040 |
| 12 | Climatic Processes/ | 106 |
| 13 | Meteorological Concepts/ | 2195 |
| 14 | precipitation.mp. | 60246 |
| 15 | Climate Change/ | 11425 |
| 16 | climate change.mp. | 20167 |
| 17 | Climate/ | 20630 |
| 18 | climat*.mp. | 82670 |
| 19 | El Nino-Southern Oscillation/ | 226 |
| 20 | el nino-southern oscillation.mp. | 520 |
| 21 | Seasons/ | 95637 |
| 22 | season*.mp. | 165185 |
| 23 | Sunlight/ | 14015 |
| 24 | sunlight.mp. | 19740 |
| 25 | Vapor Pressure/ | 347 |
| 26 | vapor pressure.mp. | 1914 |
| 27 | Atmospheric Pressure/ | 7469 |
| 28 | atmospheric pressure.mp. | 12344 |
| 29 | 1 or 2 or 3 or 4 or 5 or 6 or 7 or 8 or 9 or 10 or 11 or 12 or 13 or 14 or 15  or 16 or 17 or 18 or 19 or 20 or 21 or 22 or 23 or 24 or 25 or 26 or 27 or 28 | 905218 |
| 30 | Models, Biological/ | 317354 |
| 31 | Models, Statistical/ | 83646 |
| 32 | Statistics as Topic/ | 89885 |
| 33 | Models, Theoretical/ | 137173 |
| 34 | model*.mp. | 2898986 |
| 35 | Linear Models/ | 73006 |
| 36 | linear model.mp. | 10397 |
| 37 | Logistic Models/ | 122871 |
| 38 | logistic model.mp. | 3489 |
| 39 | Risk Assessment/ | 227411 |
| 40 | risk assessment.mp. | 247914 |
| **S1 Table**. Search strategy for OVID Medline, as performed in July 2021. | | |

**S1 Table 1**
